# Effect of disclosing a polygenic risk score for coronary heart disease on adverse cardiovascular events: 10-year follow-up of the MI-GENES randomized clinical trial

**DOI:** 10.1101/2024.07.19.24310709

**Authors:** Mohammadreza Naderian, Marwan E. Hamed, Ali A. Vaseem, Kristjan Norland, Ozan Dikilitas, Azin Teymourzadeh, Kent R. Bailey, Iftikhar J. Kullo

## Abstract

**Background:** The MI-GENES clinical trial (NCT01936675), in which participants at intermediate risk of coronary heart disease (CHD) were randomized to receive a Framingham risk score (FRS_g_, n=103), or an integrated risk score (IRS_g_, n=104) that additionally included a polygenic risk score (PRS), demonstrated that after 6 months, participants randomized to IRS_g_ had higher statin initiation and lower low-density lipoprotein cholesterol (LDL-C).

**Objectives:** In a post hoc 10-year follow-up analysis of the MI-GENES trial, we investigated whether disclosure of a PRS for CHD was associated with a reduction in adverse cardiovascular events.

**Methods:** Participants were followed from randomization beginning in October 2013 until September 2023 to ascertain adverse cardiovascular events, testing for CHD, and changes in risk factors, by blinded review of electronic health records. The primary outcome was the time from randomization to the occurrence of the first major adverse cardiovascular event (MACE), defined as cardiovascular death, non-fatal myocardial infarction, coronary revascularization, and non-fatal stroke. Statistical analyses were conducted using Cox proportional hazards regression and linear mixed-effects models.

**Results:** We followed all 203 participants who completed the MI-GENES trial, 100 in FRS_g_ and 103 in IRS_g_ (mean age at the end of follow-up: 68.2±5.2, 48% male). During a median follow-up of 9.5 years, 9 MACEs occurred in FRS_g_ and 2 in IRS_g_ (hazard ratio (HR), 0.20; 95% confidence interval (CI), 0.04 to 0.94; *P*=0.042). In FRS_g_, 47 (47%) underwent at least one test for CHD, compared to 30 (29%) in IRS_g_ (HR, 0.51; 95% CI, 0.32 to 0.81; *P*=0.004). IRS_g_ participants had a longer duration of statin therapy during the first four years post-randomization and a greater reduction in LDL-C for up to 3 years post-randomization. No significant differences between the two groups were observed for hemoglobin A1C, systolic and diastolic blood pressures, weight, and smoking cessation rate during follow-up.

**Conclusions:** The disclosure of an IRS that included a PRS to individuals at intermediate risk for CHD was associated with a lower incidence of MACE after a decade of follow-up, likely due to a higher rate of initiation and longer duration of statin therapy, leading to lower LDL-C levels.

## Introduction

Genome-wide association analyses have identified numerous loci associated with coronary heart disease (CHD).^1–3^ The aggregation of risk alleles at these loci into a polygenic risk score (PRS) provides a quantitative measure of genetic susceptibility,^4–7^ which can be integrated with other clinical and lifestyle risk factors to provide a more accurate estimate of an individual’s risk.^5,8–13^ There is great interest in using PRS for CHD in the clinical setting but little data is available regarding whether such use reduces adverse cardiovascular events.^14^ Prior observational studies suggest that individuals at high polygenic risk for CHD have greater benefit from preventive measures than individuals at low or intermediate polygenic risk.^15–17^

The MI-GENES trial (NCT01936675)^18,19^ was a randomized clinical trial (RCT) designed to assess the impact of disclosing a PRS for CHD, in addition to a conventional risk score based on the Framingham equation,^20^ on low-density lipoprotein cholesterol (LDL-C) levels. The design of the MI-GENES trial and the results have been previously published.^18,19^ The study recruited and randomized 207 non-Hispanic white participants (age 45-75 years, mean age at randomization: 59.4±5.1 years, 47% male) from Olmsted County, Minnesota, without known atherosclerotic cardiovascular disease (ASCVD), at intermediate risk for CHD (10-year risk: 5-20%), and not on statin treatment. Participants were randomized into a Framingham risk score group (FRS_g_, n=103; receiving their 10-year risk of CHD based on Framingham risk score), and an integrated risk score group (IRS_g_, n=104; who received PRS information in addition to the Framingham risk score). A genetic counselor disclosed the risk, followed by shared decision-making regarding statin therapy with a physician. Six months after randomization, participants in FRS_g_ also received their PRS for CHD via mailed letters.

Conducting a RCT to assess the long-term impact of PRS-guided prevention on adverse cardiovascular events is challenging because of the logistical burden and cost of such a study. To begin to address this knowledge gap, we undertook a 10-year follow-up study of the MI-GENES cohort to investigate whether disclosure of IRS was associated with a lower adverse cardiovascular event rate than disclosure of FRS alone and whether any differences in adverse cardiovascular events could be explained by changes in risk factors during follow-up. The long-term follow-up of the MI-GENES cohort was feasible because participants were residents of Olmsted County, Minnesota and obtained care at Mayo Clinic Rochester or the Mayo Clinic Health System.

## Methods

### Data Source

The follow-up duration in our study spanned from the beginning of randomization in October 2013 until the end of September 2023. The electronic health record (EHR) data from the Mayo Clinic Health System was reviewed to ascertain adverse cardiovascular events, testing for CHD, and changes in CHD risk factors during the follow-up period. We followed the “REporting of studies Conducted using Observational Routinely-collected Data” (RECORD) guideline for standardized reporting of data in the MI-GENES follow-up study (eTable 1 in Supplement).^21^ We updated some of the terms used in the original trial to align with current standards (eTable 2 in Supplement). The protocol for the follow-up of MI-GENES participants was approved by the Institutional Review Board of Mayo Clinic, Rochester, Minnesota (IRB#: 23-004831).

### Outcome Definition

#### Ascertainment and adjudication of major adverse cardiovascular event

The primary outcome of the follow-up study was defined as the time from randomization in the MI-GENES trial to the occurrence of the first adjudicated major adverse cardiovascular event (MACE), defined as cardiovascular death, non-fatal myocardial infarction (MI), coronary revascularization, and non-fatal stroke. Detailed definitions for each component of MACE are provided in eMethod 1 in Supplement.

Any possible MACE within the defined follow-up timeframe was initially ascertained by a manual EHR data review by two of the authors (MN, MEH), blinded to the initial randomization of participants (Figure 1). To harmonize the manual chart review process, each author first independently reviewed EHR data for 20 distinct participants and compared findings to resolve any conflicts. Following this initial phase, both physicians independently reviewed EHR data of each enrolled participant, ensuring consistency in case ascertainment.

**Figure 1:**
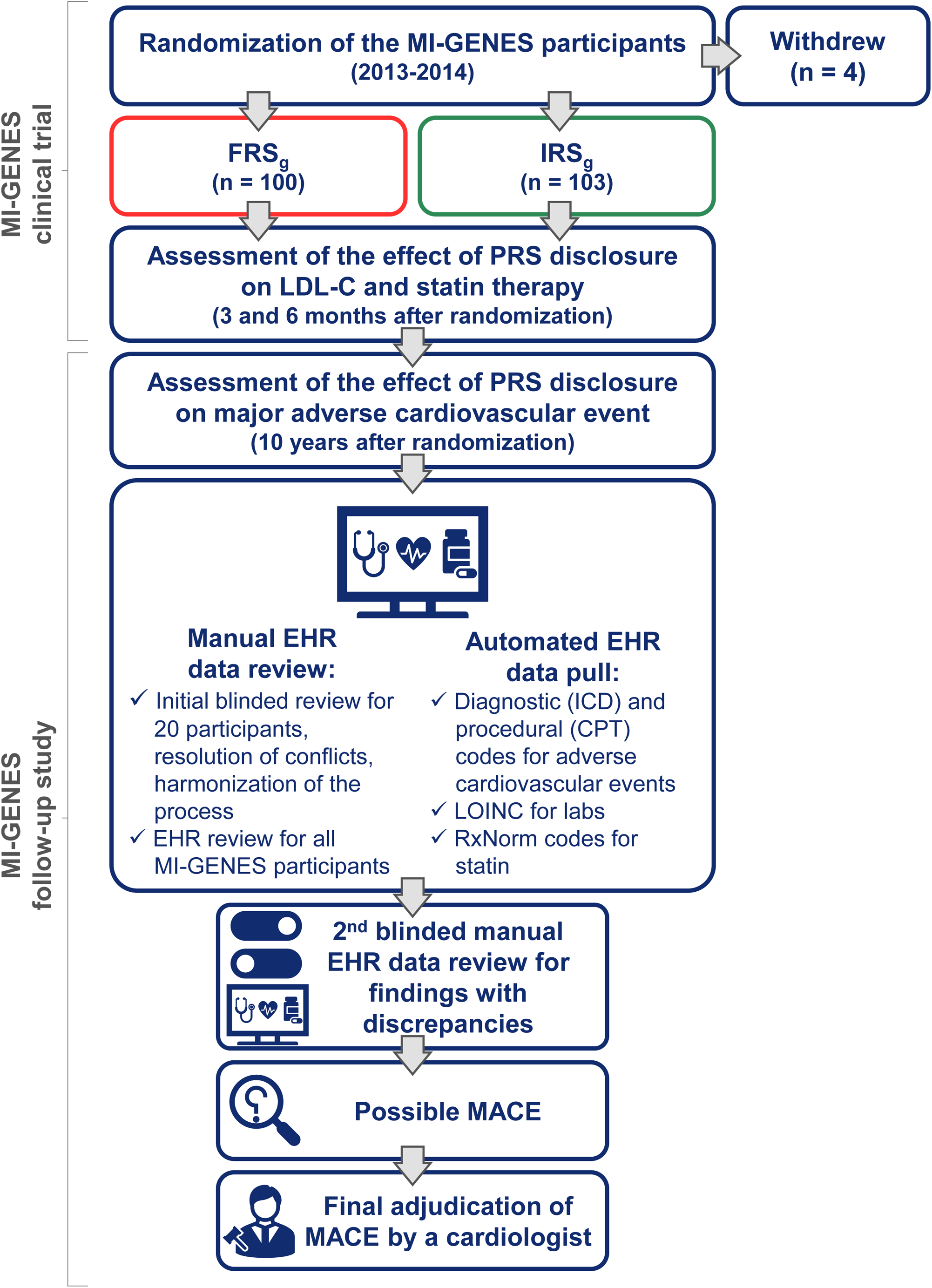
Flow Diagram of the MI-GENES Clinical Trial and the MI-GENES Follow-up Study Abbreviations: CPT: Current Procedural Terminology, EHR: electronic health records, FRS_g_: Framingham risk score group, ICD: International Classification of Diseases, IRS_g_: integrated risk score group, LDL-C: low-density lipoprotein cholesterol, LOINC: Logical Observation Identifiers Names and Codes, MACE: major adverse cardiovascular event, PRS: polygenic risk score

In addition to manual EHR data review, we also ascertained cardiovascular events by automated EHR data pull, using a set of International Classification of Diseases (ICD-9 and ICD-10) codes related to adverse cardiovascular events, as well as Current Procedural Terminology (CPT) codes for event-related procedures (detailed information on the ICD and CPT codes is presented in eTable 3 and eTable 4 in Supplement). After comparing the MACE identified through manual EHR data review with those detected through automated EHR data pull, a 2^nd^ round of manual EHR data review was conducted if the findings were discordant. Subsequently, the identified possible MACE underwent a blinded adjudication process. This multi-step approach ensured the accuracy and reliability of the extracted data for case adjudication and analysis.

For each MACE, a comprehensive review of the participant’s medical history, symptoms, physical examination findings at the time of presentation, diagnostic tests during clinical assessment, and final diagnosis, along with subsequent medical treatment was undertaken. All possible MACE underwent final adjudication by a cardiologist, who was also blinded to the initial randomization. The final adjudication of each MACE was based on predefined criteria (eMethod 1 in Supplement). In instances where multiple possible MACE were reported for a participant, each event was presented separately to the adjudicator, and the earliest definite event was considered as the MACE for that participant.

#### Stress testing and coronary artery imaging

In addition to assessing differences in MACE, we investigated the time from randomization to the first instance of testing for the diagnosis of CHD with or without non-fatal MI or coronary revascularization (eMethod 1 in Supplement). These tests for CHD encompassed stress testing and coronary artery imaging. To accomplish this, we again conducted a blinded manual EHR data review and complementary automated EHR data pull using CPT codes (eTable 4 in Supplement).

#### CHD risk factor comparison

In addition to MACE, stress testing, and coronary artery imaging, we assessed differences in CHD risk factors as a potential explanation for any observed differences in MACE. We used Logical Observation Identifiers Names and Codes (LOINC) to extract data on lipid profiles and hemoglobin A1C measurements from EHR (detailed information on LOINC in eTable 5 in Supplement). We collected data for lipid profiles from the 3-month mark after baseline and excluded post-MACE lipid profiles to only capture the effect of statins in the primary prevention setting, avoiding the confounding effect of low LDL-C and non-high-density lipoprotein cholesterol (non-HDL-C) levels attributable to statin use for secondary prevention. Additionally, we obtained available data on systolic and diastolic blood pressures (SBP and DBP), weight, and smoking cessation during the defined follow-up period.

#### Statin use

Utilizing RxNorm codes for statins, we extracted all the statin orders for MI-GENES participants to analyze the trend and differences in statin therapy between two groups (detailed information on the RxNorm codes is in eTable 6 in Supplement). To assess statin intensification and de-escalation over the follow-up period, we used accepted definitions of statin intensity.^22^ We used the start and end dates of each statin order to determine the number of participants on statin therapy at various time points during follow-up.

#### Statistical analyses

The primary outcome – time from randomization to the first MACE – was assessed using the Cox proportional hazards model. We also used Cox model to analyze time to testing for CHD. Given the randomized design of the original MI-GENES trial, Cox models were not further adjusted for other potential confounding covariates. Linear mixed-effects models were fitted to compare covariates with multiple measurements during the follow-up period between randomization groups. These covariates included LDL-C, non-HDL-C, triglyceride, hemoglobin A1C, SBP, DBP, and weight. The fixed effects included in the model were the group randomization, time difference from randomization, and their interaction, along with the baseline measurement of the covariate. Additionally, we included each participant as a random intercept to capture individual variability in multiple measured covariates. We also assessed and confirmed that the assumptions of the linear mixed-effects model were met. To compare discrete variables between the two groups, we used the chi-squared test and Fisher’s exact test. Wilcoxon’s rank test and t-test were applied to compare non-normally distributed continuous covariates and normally distributed covariates, respectively. All tests were two-sided, and statistical significance was determined at *P*<0.05. All statistical analyses were performed using R (version 4.3.1, R Foundation for Statistical Computing).

## Results

### Participants characteristics

Four enrolled participants withdrew from the MI-GENES clinical trial after randomization (3 in FRS_g_ and 1 in IRS_g_). The remaining 203 participants (100 in FRS_g_ and 103 in IRS_g_) were followed up. The mean age at the end of follow-up was 68.2±5.2 years. There was no loss to follow-up, and the duration of follow-up was similar between two groups. The last residential address for 95.6% of participants was in the state of Minnesota, similar in both groups (eTable 7 in Supplement).

### MACE during follow-up

During the follow-up period, there was 1 (1%) cardiovascular death and 4 (4%) non-fatal strokes in FRS_g_, with no corresponding events in IRS_g_. Non-fatal MI occurred in 4 (4%) FRS_g_ participants and 1 (1%) IRS_g_ participants. There was one coronary revascularization (percutaneous coronary intervention) which occurred in IRS_g_ (eTable 8 in Supplement). In total, 9 (9%) MACE occurred in FRS_g_ and 2 (2%) MACE occurred in IRS_g_ (hazard ratio (HR), 0.20; 95% confidence interval (CI), 0.04 to 0.94; *P*=0.042) (Figure 2). Based on the rate of incident MACE in FRS_g_ and IRS_g_, 14 individuals (95% CI, 0 to 53) need to be disclosed their risk with a PRS to prevent one MACE over 10 years (eTable 9 in Supplement).

**Figure 2:**
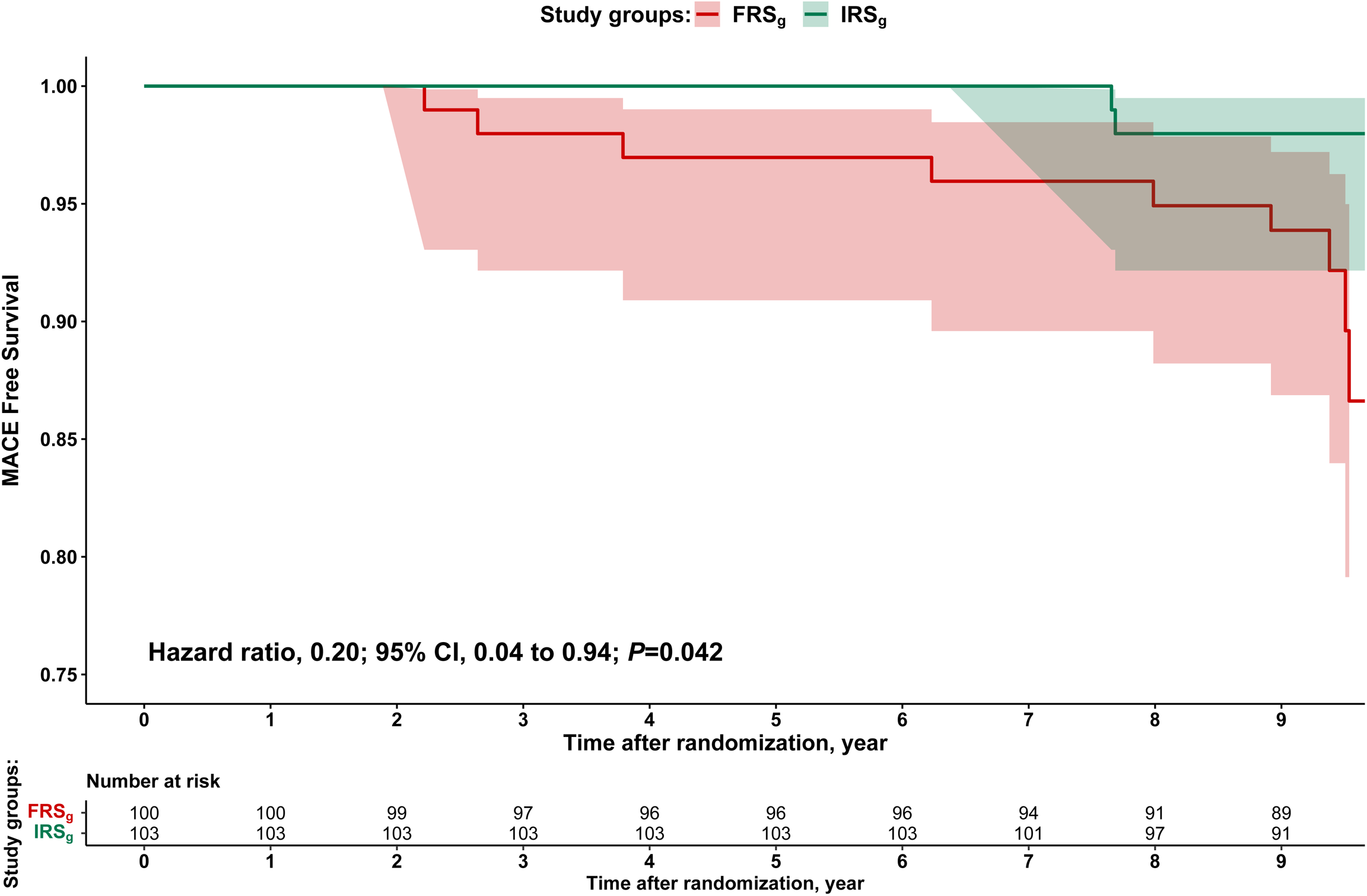
Kaplan-Meier Graph Comparing MACE Free Survival in FRS_g_ and IRS_g_ over the Follow-up Period Abbreviations: CI: confidence interval, FRS_g_: Framingham risk score group, IRS_g_: integrated risk score group, HR: hazard ratio, MACE: major adverse cardiovascular event

### Testing for CHD during follow-up

In FRS_g_, 47 (47%) underwent at least one test for CHD, compared to 30 (29%) in IRS_g_ (*P*=0.009). The most common tests were stress echocardiography, followed by coronary artery calcium scanning, and exercise ECG. The Cox model showed a HR of 0.51 (95% CI, 0.32 to 0.81) for cumulative proportion of participants in IRS_g_ compared to FRS_g_ who underwent testing for CHD (*P*=0.004) (Table 1, eFigure 1, eTable 10 in Supplement). Of participants who underwent stress testing during the follow-up period, 7 (15%) in FRS_g_ and none of the IRS_g_ participants were detected to have myocardial ischemia (*P*=0.037) (Table 1, eTable 10 in Supplement). FRS_g_ participants tended to undergo coronary angiography more often than IRS_g_ (9 (9%) vs. 3 (3%); *P*=0.079). Of the 12 coronary angiography procedures performed, 6 (50%) were reported as non-obstructive, 3 (25%) as single-vessel disease, and 2 (17%) and 1 (8%) as two-vessel disease and left-main-three-vessel disease, respectively (Table 1, eTable 10 in Supplement).

**Table 1:** Stress Testing and Coronary Artery Imaging During Follow-Up of MI-GENES Participants.

|  | **FRS<sub>g</sub>**<br>**(n=100)** | **IRS<sub>g</sub>**<br>**(n=103)** |
| --- | --- | --- |
| Participants with at least one test for CHD during follow-up<sup>a</sup> | 47 (47%) | 30 (29%) |
| Exercise ECG | 13 | 10 |
| Nuclear perfusion imaging | 5 | 6 |
| Stress echocardiography | 23 | 11 |
| CAC scan | 18 | 11 |
| Coronary CT angiography | 4 | 7 |
| Coronary angiography | 9 | 3 |
| Participants with proven ischemia<br>on a stress test<sup>b, c</sup> | 7 (15%) | 0 (0%) |

<sup>a</sup> The numbers might overlap for testings for CHD as some participants underwent >1 stress testing or coronary artery imaging.
<sup>b</sup> Percentages have been reported based on the number of participants with at least one testing for CHD during follow-up.
<sup>c</sup> For some participants who tested positive in stress testing (exercise ECG, nuclear perfusion imaging, and stress echocardiography) subsequent coronary artery imaging via coronary CT angiography or coronary angiography did not indicate ischemia.
Abbreviation: CAC: Coronary Artery Calcium, CT: Computed Tomography, ECG: Electrocardiogram, FRS<sub>g</sub>: Framingham risk score group, IRS<sub>g</sub>: integrated risk score group

### Lipid profiles during follow-up

The frequency of lipid profile measurements during the follow-up period was similar between two groups (eTable 7 in Supplement). As reported in the original MI-GENES paper, LDL-C levels were significantly lower in IRS_g_ at 6 months post-randomization and in our follow-up analyses, the differences remained significant for up to 3 years (Figure 3 - top panel, eTable 11 in Supplement). Additionally, individuals in IRS_g_ showed greater reductions in non-HDL-C and triglyceride levels compared to those in the FRS_g_ for up to 4-and 2.5-years post-randomization, respectively (Figure 3 - middle and bottom panels, eTable 11 in Supplement).

**Figure 3:**
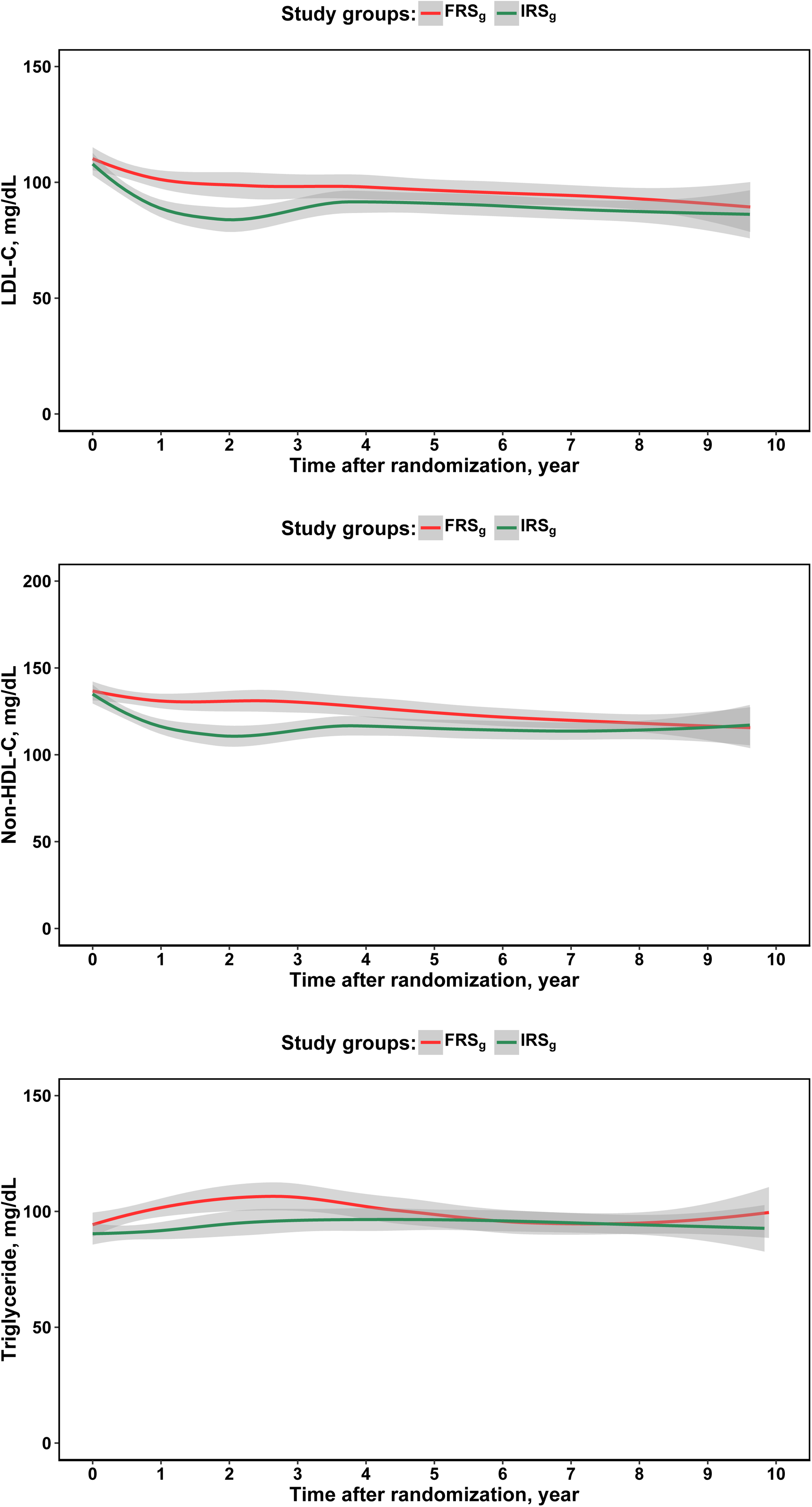
Predicted LDL-C, Non-HDL-C, and Triglyceride Levels Over 10 Years of Follow-up Predicted mean LDL-C (top panel), non-HDL-C (middle panel), and triglyceride (bottom panel) levels from randomization to the end of follow-up period, utilizing a locally weighted scatterplot smoothing (LOESS) model on actual data. The solid lines represent the predicted mean values, while the shaded regions indicate the corresponding 95% confidence intervals. Estimated differences for these measurements using linear mixed-effects models are provided in eTable 11 in Supplement. Abbreviations: FRS_g_: Framingham risk score group, IRS_g_: integrated risk score group, LDL-C: low-density lipoprotein cholesterol, Non-HDL-C: non-high-density lipoprotein cholesterol

### Statin therapy during follow-up

During the initial four years post-randomization, a higher proportion of participants in IRS_g_ were taking statins than in FRS_g_. This proportion gradually declined and eventually reached a level similar to that of FRS_g_ four years after randomization (Figure 4, eFigure 2 in Supplement). No significant differences were observed between the two groups regarding low and high-intensity statins; however, during the initial four years post-randomization, a higher proportion of participants in IRS_g_ were taking medium-intensity statins than in FRS_g_ (Figure 4, eFigure 2 in Supplement).

**Figure 4:**
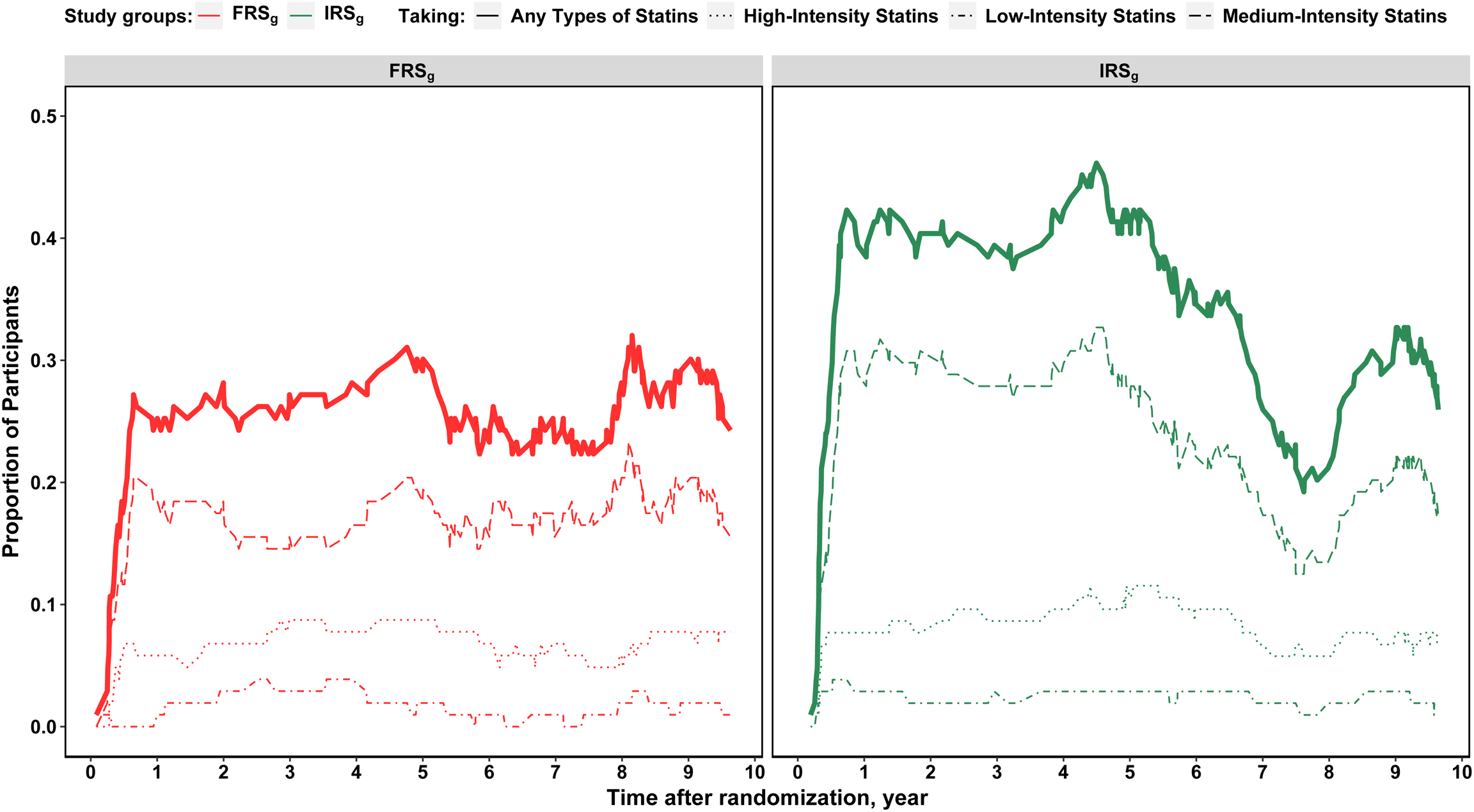
Proportion of Participants on Statin Therapy in FRS_g_ and IRS_g_ over 10 Years of Follow-up This figure shows the proportion of participants in FRS_g_ (left panel) and IRS_g_ (right panel) who were taking low-intensity (dotted dashed lines), medium-intensity (dashed lines), high-intensity (dotted lines), and any types of statins (thick solid lines), during 10-year follow-up period. Abbreviations: FRS_g_: Framingham risk score group, IRS_g_: integrated risk score group

### Trajectories of other CHD risk factors during follow-up

The frequency of hemoglobin A1C, SBP, DBP and weight measurements during the follow-up period was similar between two groups (eTable 7 in Supplement). No significant differences were observed in mean hemoglobin A1C levels, mean SBP and DBP levels, and mean weight between the two groups throughout the follow-up period (eFigure 3, eTable 12 in Supplement). Three of the six participants in the IRS_g_ identified as current smokers at randomization, quit smoking during the follow-up period, whereas among the six participants in the FRS_g_ identified as current smokers at randomization, only one quit smoking (eFigure 4 in Supplement).

## Discussion

In this 10-year follow-up study of the MI-GENES clinical trial, we observed a lower incidence of MACE (cardiovascular death, non-fatal MI, coronary revascularization, and non-fatal stroke) in those randomized to disclosure of an IRS (comprising both PRS and FRS) vs. FRS alone. Additionally, the disclosure of IRS was associated with a lower rate of testing for CHD, suggesting lower incidence of CHD symptoms. In the original report of the MI-GENES clinical trial, disclosure of IRS for CHD compared to FRS alone, led to a higher rate of statin therapy and reductions in LDL-C levels, at 6-months following disclosure. During the 10-y follow-up, individuals who received IRS were on statin for a longer duration, leading to more durable reductions in LDL-C levels than in the control group (FRS_g_). A lower cumulative LDL-C exposure due to earlier statin initiation and longer duration of statin therapy could be one reason for the lower rate of MACE in IRS_g_.

The MI-GENES cohort was relatively small and limited to participants at intermediate risk for CHD (10-y risk: 5-20%) resulting in a modest number of MACE. The differences in MACE, mean LDL-C levels, and statin use duration were significant, but should be interpreted with caution given the modest sample size and the limited number of events, which is also reflected in the relatively wide confidence intervals. One cannot exclude the possibility that the results are due to chance. However, lower cumulative LDL-C exposure and lower rate of new CHD symptoms and detectable ischemia in stress testing in the IRS_g_ increase confidence that the finding of lower MACE in IRS_g_ is a true effect. Larger clinical trials, nonetheless, are needed to confirm these findings.

Cumulative LDL-C exposure over time is associated with increase in cardiovascular risk, underscoring the importance of earlier LDL-C control for prevention.^23^ In several clinical trials, earlier initiation of statins, the use of higher statin intensity, and more prolonged duration of statin therapy were associated with improved cardiovascular outcomes i.e. a ‘legacy effect’.^24–26^ The significantly higher proportion of statin prescription and therapy during the follow-up period in the IRS_g_ was mostly due to greater use of medium intensity statins. The differences, however, were noted only in the initial half of the follow-up. In part this was due to later discontinuation of statins possibly because of perceived adequate LDL-C reduction in the primary prevention setting or higher reported side effects by IRS_g_ participants (Figure 4, eFigure 2 in Supplement).

An analysis of the motivation, perception, and treatment beliefs of the MI-GENES participants indicated that those who received IRS at the time of randomization had a stronger desire to improve their health.^27^ Additionally, they had higher perceived personal control, reflecting their belief in being able to alter their situation by bringing about desirable change through cognitive, behavioral, and decisional control.^28^ While we were not able to capture longitudinal data on lifestyle modifications to assess long-term behavioral changes between the FRS_g_ and IRS_g_, difference in perceived personal control shortly post-randomization raises the possibility that some of the difference in MACE between the two groups could have resulted from favorable lifestyle modifications in IRS_g_ during the follow up period. Disclosure of an IRS was also associated with increased information seeking and information sharing behaviors, particularly with family members, friends, coworkers, and primary care providers.^29^ Such behavior may have led to increased knowledge about risk factors for CHD and motivation to control these as well as seek continuity of care with their care provider.^29^

### Strengths and limitations

This follow-up study of the MI-GENES trial participants provides for the first-time data on long-term outcomes of PRS-guided prevention for ASCVD. Remarkably, follow-up was possible in >95% of the initially enrolled participants, primarily because they continued to reside in Minnesota and receive care at the Mayo Clinic or one of the regional Mayo Clinic Health System centers. Thus we had access to comprehensive data with minimal loss to follow-up over 10 years. In addition, we performed a multistep blinded process for both manual and automated data extraction from EHR to ensure accurate ascertainment of MACE and testing for CHD.

We acknowledge several limitations of our study, including a relatively small sample size limited to participants at intermediate risk for CHD (5-20%) resulting in a modest number of incident MACE. Because the MI-GENES trial was designed before availability of the pooled cohort equations,^30^ the Framingham risk score was used to estimate 10-y CHD risk. The PRS utilized in the MI-GENES trial was computed from 28 SNVs. As of 2013, ∼ 50 SNVs had been identified as genome-wide significant, and 22 SNVs in risk factor pathways (lipid, blood pressure, and diabetes mellitus) were excluded from the PRS used in the MI-GENES trial. Additionally, lack of diversity in the study cohort limits the generalizability of the findings to other racial/ethnic groups. The study was conducted at an academic medical center, CHD risk was disclosed by a genetic counselor followed by shared decision-making with physicians in the Cardiovascular Health Clinic. Such implementation may be challenging to replicate in primary care practice and low resource centers and additional studies are needed in diverse settings in the United States.

## Conclusion

In this 10-year follow-up study of the MI-GENES trial, disclosure of an integrated CHD risk score that included a PRS, to individuals at intermediate risk of CHD, was associated with a lower incidence of MACE. A higher rate of early initiation of statins, longer duration of statin therapy and lower LDL-C levels during the first four years after disclosure of an integrated CHD risk, likely contributed to this effect. In addition, those receiving an integrated CHD risk underwent testing for CHD less frequently, suggestive of a lower incidence of CHD symptoms. The results of this study motivate larger RCTs that enroll from diverse practice settings to confirm our findings.

## Data Availability Statement

The dataset generated and analyzed for the current study is available from the corresponding author on reasonable request that might be necessary to interpret, replicate, and build upon the methods or findings reported.

